# Strategies and Tools for electronic health records and physician workflow alignment: A scoping review protocol

**DOI:** 10.1101/2023.12.27.23300587

**Authors:** Oluwakemi O. Oluwole, Nicole Haggarty, Uche Ikenyei, Oluwabambi Tinuoye, Andreawan Honora, Mohammed Abass Issakah

## Abstract

**Introduction:** The rapid adoption of electronic health records (EHR) across the globe by the healthcare industry is an indication of the rising digitalization of healthcare functions. Despite the potential benefits of EHR, achieving a fit between physician workflow and EHR has posed a major challenge, with negative effects on physician wellbeing and patient outcomes. To this effect, organizations have attempted to align the EHR with physician workflow in various ways. It is important to understand the strategies and tools that have been employed and, where possible, the outcome of these engagements as a fundamental insight to resolving this issue.

**Methodology:** The study will employ a methodological framework developed by Arksey and O’Malley to strategically identify articles that use any type of concept for the alignment of physician workflow and EHR from Embase (OVID), MEDLINE (OVID), PubMed, CINAHL, and Scopus databases. It will follow the Preferred Reporting Items for Systematic Reviews and Meta-Analysis Extension for Scoping Reviews (PRISMA-ScR) checklist to report findings. The articles will be extracted into the Covidence software, screened, and relevant data will be extracted from the selected articles. A qualitative thematic approach will be used to analyze the data. No ethical approval was sought because the data were collected from sources in the public domain.

**Result:** The scoping review is scheduled to be completed by April 2024. The results will be presented in tabular and narrative form and published in a reputable journal and through conference presentations.

**Conclusion:** The outcome of the review aims to provide a systematic toolkit of activities and strategies and, where possible, the corresponding effectiveness that organizations can use to optimize EHR-to-physician workflow alignment. The outcome would also help make appropriate recommendations for alignment. Subsequently, it can improve patient outcomes such as the reduction of medication errors and improvement of patient-centered, improve physician well-being and reduce burnout.

## Introduction

The adoption of electronic health records (EHRs) is increasing across national and international healthcare systems [1]. EHRs are rapidly becoming an acceptable means of digitalizing health data, making it more accessible to patients, and improving quality and analytical opportunities that improve patient outcomes [2, 3]. Notwithstanding the acknowledged potential of the EHR to improve healthcare, the claim that it improves workflow efficiency and productivity appears to be overstated.

A workflow is a collection of activities organized sequentially into processes; it also includes the resources required to complete the task [4]. A workflow is a series of steps linked together. It is different from a task because it is based on the flow paradigm. A workflow is a model that represents the steps involved in a real work environment to achieve a specified goal. It is used to map out the sequential tasks required in taking a work description from ‘initiated’ to ’processed’. For example, steps involved in patient-physician interaction from a patient’s arrival at the physician’s office to their departure from the office can be captured as a workflow [5]. Oftentimes, workflow design represents a consistent sequence of tasks involved in the operation. However, the complex and emergent nature of healthcare and variation in treatment processes sometimes conflict with the use of standardized processes in the EHR [5], which generates workflow challenges. While the duties of some health professionals, such as nurses, lean more toward standard routines, the duties of physicians vary greatly with each physician and patient [6]. Using a standard EHR workflow design for physicians can be difficult.

Several physician workflow challenges have also been documented in the EHR literature [7, 8] For example, some authors have mentioned that EHR designers who are non-healthcare professionals are sometimes unaware of the need to make EHRs adaptable, flexible, and responsive to the unpredictable nature of routine physician workflows [5, 9]. EHR designers may intend to increase the value of EHRs at the expense of physicians’ clinical routines and workflow, resulting in usability obstacles. EHRs are not designed to automate physicians’ exact paper-based workflows [5]. The corresponding effects of workflow interference on physicians are unpleasant and have been recognized as a significant contributor to physician burnout [10, 7, 8]. Furthermore, physician workflow design in the EHR can fail to support foundational physician-patient interaction, limit assessment and treatment, and increase the risk of medication errors [11]. Suboptimal EHR-mediated workflows can inform EHR evasion, leading to workarounds. Workarounds are other ways of using EHRs other than the recommended methods after system implementation [12]. This mismatch is often associated with physician stress [13], which can result in inadequate and erroneous patient documentation [4]. Given that physician workflow is an important determinant of maximizing the benefits of the EHR [14], understanding the EHR-physician workflow alignment is of prime importance to achieving successful EHR implementation and use.

Despite the rapid growth and proliferation of studies on the development of strategies and tools to improve alignment between physicians and EHR workflows, the existing knowledge is highly heterogeneous and fragmented, presenting gaps that hinder a comprehensive and holistic view. We argue that to advance our understanding of how EHR-physician workflow alignment should be performed, it is essential to map the intellectual structure of current knowledge. This would be achieved by synthesizing the concepts in all publications that focus on EHR-physician workflow alignment through a scoping review. Therefore, the aim of this study is to identify the strategies and tools that organizations have used to optimize physician workflow and to create a systematic toolkit of concepts and activities to achieve maximum alignment and minimize obstacles to physician workflow in EHR implementations. This study will provide an overview and assessment of the tools and strategies used in the literature to build a systematic toolkit and identify potential research gaps. This protocol explains the objectives, plans, and steps that reviewers will take to extract data, analyze it, and report the results of the review.

### Search for similar scoping reviews

Before starting a scoping review, it is important that the literature be searched for similar topics and goals. We found a study by Avendano et al. that compared three randomly chosen EHR-physician alignment strategies instead of systematically elaborating all of them [15]. Our review intends to identify all strategies and tools used by organizations to align the EHR with physician workflow. Also, study by Kruse et al. explored the effect of organizational directed workplace intervention on physician burnout [16]. Although physician workflow challenges have been associated with EHR, the focus of our review is on the technology, and how organizations have improved its integration to physician workflow. In August 2023, searches in the Cochrane and JBI databases did not reveal similar scoping review protocols or title registrations. A scoping review was chosen to synthesize the strategies and tools used to align EHR and physician workflow, presenting diverse research innovations, their impacts and to identify potential research gaps.

## Methods

A scoping review method was carefully chosen to synthesize different types of evidence across databases from published literature [17]. The scoping review was also necessary to map the fundamental concepts to a research area and to elucidate related definitions. Therefore, this protocol is a proposal for a review of the existing literature to provide evidence on workflow optimization by aligning EHRs with physician workflow. The review’s goal is to identify the strategies and tools that organizations have employed in an effort to harmonize physician workflow with the EHR. In addition, this scoping review will provide an overview of the effectiveness of these strategies on EHR physician workflow.

Furthermore, the review is expected to provide insight into the gaps in the effort to align the EHR with physician workflow and provide a direction for further studies on the subject. To maximize rigor, this study will follow the guidelines provided by Arksey and O’Malley’s framework for scoping review [17]. The guideline provides the following steps: identifying the research question for the study, identifying studies relevant to the research question, study selection, data charting, result collation, summarization, and reporting. All members of the research team designed, reviewed, and approved this protocol. The Preferred Reporting Items for Systematic Reviews and Meta-Analyses Extension for Scoping Reviews (PRISMA-ScR) checklist informed the development of the protocol. This will enhance the completeness of the report and provide transparency [18].

### Step 1: Formulate a research question

This study seeks to address one key question; what strategies and tools are used by organization in the attempt to align EHR to physician workflow?

#### Population of the Study

The targeted population for this study was physicians. Article selection will be limited to those with physicians who have direct interaction with patients. Non-patient-engaging physicians, such as pathologists, will be excluded from the study because they interact with the EHR in a different way and face different challenges than physicians, who need to combine documentation and review with patient interaction. This study will consider articles that focus on physicians within outpatient or inpatient hospitals and primary health care settings. In addition, Peters et al. argued that the focus of the study should be established by indicating the population, concept, and context (PCC) [19]. Table 1 shows the PCC of the study.

**Table 1:** Population, context and concept (PCC) adopted in the study.

|  |  |
| --- | --- |
| Population | Physicians |
| Concept | EHR to physician workflow alignment. |
| Context | Workflow of physicians that have direct contact with patients in any context (in-patient, out-patient, hospitals, and primary health care) will be included. |

#### Concept

The concept explored in this review is the alignment of EHR with physician workflow. Although many authors do not refer to this concept as alignment, the phrases ‘EHR workflow optimization’ and ‘workflow optimization’ are often used. However, this study will focus on EHR optimization, which seeks to align physician workflow with EHR design.

#### Context

The context of physician workflow considered in this review is based on the EHR. All studies that do not consider physician workflow with respect to the EHR will be excluded. The context of the study will not be limited to physicians working in hospitals or primary care physicians because the objective of the study is to understand the concepts and methodologies that have been applied to align physicians’ workflow with EHRs and their effectiveness. Furthermore, studies that are based on electronic medical records but meet the identified inclusion criteria for EHRs will be included. This is because the study is not concerned with the complexity of the information system in terms of its network or interconnectivity but with the improvement of physicians’ experience of the EHRs in terms of their workflow. There will be no restrictions on the search strategy in terms of year or research design, but it will be limited to publications disseminated in English to reduce translation challenges.

Although the workflow of physicians in primary healthcare, hospital settings, and different specialties (such as surgery and pediatrics) may differ, the goal of this study is to provide a broad understanding of the subject. Therefore, all studies conducted in hospital environments, primary health care, the public and private sectors, and all areas of specialization such as emergency medicine, general practice, and surgery will be included without bias for countries or continents.

### Step 2: Identification of relevant studies

To achieve this goal, both free text and controlled terms will be used and refined repeatedly. The study will search the following databases for published articles: Embase (OVID), MEDLINE (OVID), <u>CINAHL</u>, PubMed, and Scopus. Published articles will be extracted on the basis of the following terms and keywords: physician workflow, EHRs/Electronic Medical Records (EMR), clinician workflow, and optimization. The ancestry method for screening the references of selected articles will be performed manually to determine if there are articles that are still relevant to the study. Abstracts of relevant articles will be retrieved and reviewed on the basis of the inclusion and exclusion criteria. The references to newly selected studies will be screened for relevant articles; this process will continue iteratively until studies relevant to the review are exhausted. Google Scholar and ProQuest databases will be searched for unpublished articles. For example, by using free text terms in the preliminary search of the PubMed database, 14 articles were returned. The sample search strategy is presented in Table 2. This strategy will be repeated for all listed databases.

**Table 2:** A sample of the search query.

| Search number | Search Query | Items found |
| --- | --- | --- |
| #1 | Search: (((((Electronic Health Records[Title/Abstract]) OR (EHR[Title/Abstract])) OR (Electronic Medical Records[Title/Abstract])) OR (EMR[Title/Abstract])) OR (Computerized physician order entry[Title/Abstract])) OR (Electronic Documentation[Title/Abstract])) | 48,182 |
| #2 | Search: ((physician workflow[Title/Abstract]) OR (clinician workflow[Title/Abstract])) OR (provider workflow[Title/Abstract]) | 214 |
| #3 | Search: ((optimization[Title/Abstract]) OR (alignment[Title/Abstract])) OR (improvement[Title/Abstract]) | 1,060,230 |
|  | #1 AND #2 AND #3 | 14 |

### Step 3: Study selection

A scoping review protocol should clarify the criteria for eligibility (inclusion) and the sources of information used in the study [18]. The selection of an article will be strictly based on whether it focuses on the optimization of the physician workflow within the EHR. Studies that look at how to make the EHR work best for physicians by introducing strategies, models, or an understanding of how to align the workflow of physicians with the EHR will be included. The primary outcome of this study includes themes on strategies, tools, and an understanding of how to align the EHR with physician workflow and their effectiveness. The review’s secondary goals will be to show how research on the subject has changed over time, to explain where research is going, and to identify gaps in the literature. Physicians will be the focus of the study, and the goal of the study will be to systematically document and describe the tools and strategies used to optimize physician workflow with the EHR. Although some studies on clinician workflow will be included in the search by including the search term "clinician workflow," this is to ensure that studies that combine physician workflow with nurses’ workflow are not missed. Subsequently, we will exclude studies based solely on nurses’ workflow during abstract or full-text screening.

Studies that do not focus on physicians but on other healthcare professionals—for example, pharmacists and laboratory technicians—that focus on workflow for purposes other than optimizing the EHR will be excluded. We will not include any changes to the workflow that do not fix the misalignment problem; therefore, functional accessories to the EHR—for example, the study by Dummett et al. that adds new software functions to the EHR workflow, even though these functions were not part of physician workflow [20]. However, software integrated to augment physicians’ tasks that were not captured well by EHRs will be included. In addition, studies that investigate the impact of EHR design on physician workflow and those that access physician workflow with a focus on meaningful use will be excluded. The objectives of meaningful use do not focus on the efficiency of EHR physician workflow and its impact on physicians, but rather on the impact of EHR on adoption and quality of care.

Data will be uploaded into the Covidence systematic review software developed by Veritas Health Innovations. This software will automatically eliminate duplicates, manage inclusion and exclusion criteria, and track reviewer activities. The Preferred Reporting Items for Systematic Reviews and Meta-Analyses Preferred Reporting Items for Systematic reviews and Meta-Analyses extension for Scoping Reviews (PRISMA-ScR) guidelines will be followed. Two independent reviewers will screen each article for eligibility. The first phase of screening will be conducted independently, with two reviewers screening each article based on the publication title and abstract based on the inclusion and exclusion criteria. A regular meeting will be scheduled to resolve disagreements between reviewers; however, when there is doubt, a more experienced investigator will resolve the difference. A consensus report will be provided to the team to facilitate conversations on eligibility agreements. In the second phase of screening, the full text of each article will be screened in pairs to determine eligibility.

### Step 4: Data extraction

Data will be extracted from eligible articles after the full-text reviews into a standardized data extraction table in Microsoft Excel format for analysis by two reviewers. Two reviewers will design the data extraction table by adding all the variables that need to be extracted from the publications. The Excel table will be standardized through the joint efforts of all reviewers, as they will have the opportunity to contribute to the spreadsheet and discuss the ideas emerging during the review process. The characteristics extracted from each article will be added to the following variables: author’s name, year of publication, setting, methodology, type of intervention, and outcome. The two reviewers will independently extract the variables from the selected articles into the standardized extraction table and independently summarize their findings. These findings will be shared among the five reviewers, and a meeting will be held to further synthesize the findings.

### Stage 5: Result collation, summary, and report

There will be no generalization of the results because the review does not attempt to validate the quality of the articles used in the analysis [17]. Using an interpretive translation of the findings, the results will be presented in a descriptive format that fits the goal of the study. The findings will be presented in a detailed and organized manner, using tables, figures, and descriptive summaries.

### Stage 6: consultation

As an important part of the scoping review method, stakeholders should be consulted. Experts in health information science will be consulted for this study. These experts know how to set up and use an EHR and how to interpret and share the results. Two stakeholders who are both experts in EHR studies and implementation will be consulted to ensure that the design, interpretation of findings, and knowledge transfer of the scoping review are relevant to physicians.

## Result

Electronic searches of the five databases listed as of October 15, 2023, produced 564 references; after removing 66 duplicates, the Covidence software left 498 articles. Title and abstract screenings have begun. Additional data will be collected by screening more relevant articles from the references of selected articles. The scoping review is scheduled to be completed by April 2024. The results will be presented in tabular and narrative form and published in a reputable journal and through conference presentations. While the utmost effort will be made to follow the guidelines of this protocol, any deviation, uncovered information, and identified limitations will be reported.

## Discussion

The results of this review will inform stakeholders in EHR: policymakers, vendors, designers, developers, healthcare managers, and physicians. We will provide a toolkit that various organizations have used to address the misalignment between the EHR and physician workflow across various healthcare organizations and countries. Also, where possible, provide the impact of each tool and strategy. Thus, healthcare organizations can improve the alignment between implemented EHRs and physician workflow. It can help them identify the pitfalls they have encountered in the past and avoid repeating these mistakes. In addition, the results will provide evidence required by vendors and developers to guide the design and development of better EHR systems with improved EHR-to-physician workflow alignment. Furthermore, we identified knowledge gaps for future improvement in the alignment between physician workflow and EHR.

## Data Availability

Data will be provided as supplemental material when findings of the scoping review is published.

